# Epstein-Barr virus influences the induction of protective antibody responses in young African children vaccinated with the RTS,S malaria vaccine

**DOI:** 10.64898/2026.11.03.26367212

**Authors:** Alexander W. Harris, Adam Thomas, Jean-Bosco Ouedraogo, Halidou Tinto, Issaka Zongo, Alassane Dicko, Issaka Sagara, Daniel Chandramohan, Brian Greenwood, Liriye Kurtovic, James G. Beeson

## Abstract

The RTS,S vaccine provides moderate protection against malaria in children, which may be due in part to environmental factors. Cytomegalovirus (CMV) and Epstein-Barr virus (EBV) infections are prevalent in Africa and potentially contribute to suboptimal vaccine responses. We evaluated whether latent CMV/EBV infections impact RTS,S-induced antibodies in 314 vaccinated children aged 5-29 months in West Africa. While no strong associations were observed for CMV, children seropositive for EBV had lower antigen-specific IgG and IgA, which have been linked with protective malaria immunity. These results may partly explain why malaria vaccines are suboptimal in populations where early CMV/EBV infections are prevalent.

## Background

Highly effective interventions would greatly aid global efforts to reduce and control *Plasmodium falciparum* malaria, which disproportionally affects young children in sub-Saharan Africa. RTS,S and R21 are the only two World Health Organization (WHO) recommended malaria vaccines and are based on the same antigenic construct of the *P. falciparum* circumsporozoite protein (CSP). These vaccines provide moderate efficacy against malaria (∼55-75%), which wanes over time and requires booster doses to sustain partial protection. Furthermore, the induction of protective antibodies is highly variable among vaccinated children for reasons that are poorly understood^1^. Identifying environmental factors that potentially impair vaccine responses is important for developing strategies to enhance vaccine effectiveness in target populations.

Cytomegalovirus (CMV) and Epstein-Barr virus (EBV) are part of the *Herpesviridae* family and establish life-long latent infections within a range of immune cell types. The prevalence of CMV and EBV in Africa is high, with most infections occurring in the first years of life and almost 100% of individuals are estimated to have a latent infection by adulthood^2,3^. These viruses have been shown to extensively modulate the host immune system^4,5^. However, the impact of latent CMV or EBV infection on vaccine-induced responses in children remains underexplored. In Gambian infants, antibody responses to measles and meningococcal vaccines were lower among EBV+ infants, whereas CMV infection had little impact^6^. In non-malaria exposed adults, vaccine-induced antibody responses to influenza and an MSP1-based malaria vaccine candidate were poorer in CMV+ individuals^7,8^, although these findings may differ in paediatric populations. The relevance of CMV and/or EBV infection for immunisation programs remains an important yet unanswered public health question. More data on the impact of CMV/EBV infection on response to vaccination in children, especially for vaccines with suboptimal efficacy and in populations where these viral infections are prevalent, are required.

We investigated associations between CMV or EBV serostatus and vaccine-induced IgG, IgA, and IgM to two major regions of the CSP vaccine antigen in young West African children who had received the primary 3-dose (n=113) or the booster 4-dose (n=201) RTS,S regimen in the course of a completed phase III clinical trial^9^.

## Methods

### Study population and ethics approval

We evaluated sera collected during a completed, individually randomised, controlled phase III clinical trial of the RTS,S/AS01_E_ vaccine (subsequently referred to as RTS,S) administered with or without seasonal malaria chemoprevention (SMC) in Burkina Faso and Mali (ClinicalTrials.gov, NCT04319380)^9^. Briefly, children aged 5-17 months at enrolment were randomly assigned to receive either RTS,S (3-dose primary regimen in 2017, followed by a fourth booster dose in 2018), SMC (4 rounds of sulfadoxine–pyrimethamine and amodiaquine during each malaria transmission season), or a combination of RTS,S with SMC. The current study evaluated samples from both RTS,S arms (with or without SMC) to investigate vaccine-induced immunity. It has been previously shown that the anti-CSP antibody response is comparable between both RTS,S arms and between study sites^10^. Therefore, samples were pooled from each group for this analysis. Due to logistical and acceptability considerations, serum samples were collected from different children in 2017 and in 2018, which we refer to as the primary (n=113) and booster (n=201) cohorts, respectively. Samples collected prior to vaccination (pre-vac; day of the first dose for the primary cohort, and day of the fourth dose for the booster cohort) were used to assess CMV/EBV serostatus. Matched samples collected one month after completing the 3-dose primary or 4-dose booster regimen (post-vac) were used to evaluate vaccine-induced antibodies. Ethics approval was obtained by the ethics committees of the London School of Hygiene & Tropical Medicine, the Health Research Ethics Committee of Burkina Faso, the Institutional Ethics Committee of IRSS in Burkina Faso and the Ethics Committee of the Faculty of Medicine, Pharmacy and Dentistry, University of Bamako and Alfred Human Research and Ethics Committee (approval number: 458/24). Written informed consent was obtained from parents/guardians of children.

### Determination of CMV and EBV serostatus

Pre-vac samples were evaluated for anti-CMV IgG and anti-EBV IgG using enzyme-linked immunosorbent assay (ELISA) kits as per manufacturer’s instructions (ab108724 and ab108730 Abcam, UK). Briefly, samples were tested at 1/100 dilution and seropositivity was defined as an optical density (OD) value greater than the manufacturer’s cut-off control + 10%.

### Vaccine-induced antibody detection by ELISA

Antibodies were measured to a synthetic peptide representing the central repeat region^11^ (NANP) and a recombinant protein representing the C-terminal region^11^ (C-term) of CSP, which both form the basis of the RTS,S vaccine. We quantified antigen-specific IgG, IgM, and IgA isotypes by ELISA as previously described^1^. Briefly, 384-well plates (PerkinElmer, USA) were coated with antigen at 0.5□µg/ml in 1 × phosphate buffered saline (PBS). The plates were blocked with 0.1% (w/v) casein (Sigma-Aldrich, USA) in 1 × PBS, and post-vac samples were tested at 1/2000 (for IgG) or 1/1000 (for IgM and IgA) dilution using the JANUS automated workstation (PerkinElmer). Antigen-specific isotypes were detected using goat anti-human IgG (1/1000; Invitrogen, USA), IgA (1/5000; Abcam), or IgM (1/2500; Sigma-Aldrich) conjugated to horseradish peroxidase. Plates were then incubated with ABTS substrate (Thermo Fisher Scientific) and OD was measured at 405□nm. Samples and detection antibodies were prepared in 0.1% casein, and plates were washed thrice in between steps using 0.05% (v/v) Tween20 in 1 × PBS.

### Statistical analysis

Raw antibody data were corrected for background reactivity using no-serum (buffer-only) controls, and a seropositivity cut-off was determined using the median antibody levels of samples in the SMC arm included as an unvaccinated control (2017, n=25; 2018, n=25). Mann-Whitney tests were performed using GraphPad Prism 10. Multiple linear regression analyses were performed using R Studio. The interpretation of differences between groups considered the p-value (p<0.05 was considered significant), the effect size, and the overall coherency of the findings.

## Results

### CMV and EBV prevalence in pre-vac samples

Pre-vac samples were evaluated to determine the CMV and EBV serostatus of each participant (Supplementary Figure 1). In the primary cohort (n=113), 77% of children were CMV+ and 40.7% were EBV+ (Table 1). CMV+ children were significantly older (P=0.0131) and more likely to be male (P=0.0212) than CMV-children. The EBV+ children were also significantly older than EBV-children (P=0.0035) but did not vary by sex (P>0.9999). In the booster cohort (n=201), seropositivity was much higher and most children were CMV+ (92.5%) or EBV+ (78.6%). Neither age (P>0.1543) nor sex (P>0.1791) was significantly different among the seropositive and seronegative groups for each virus. We also explored associations in children who were double-positive and double-negative for both viruses and found similar trends regarding age and sex (Supplementary Table 1). The serostatus of CMV and EBV were comparable between study sites for both the primary and booster cohorts (P>0.0983 for all tests; Supplementary Table 2).

**Table 1.**
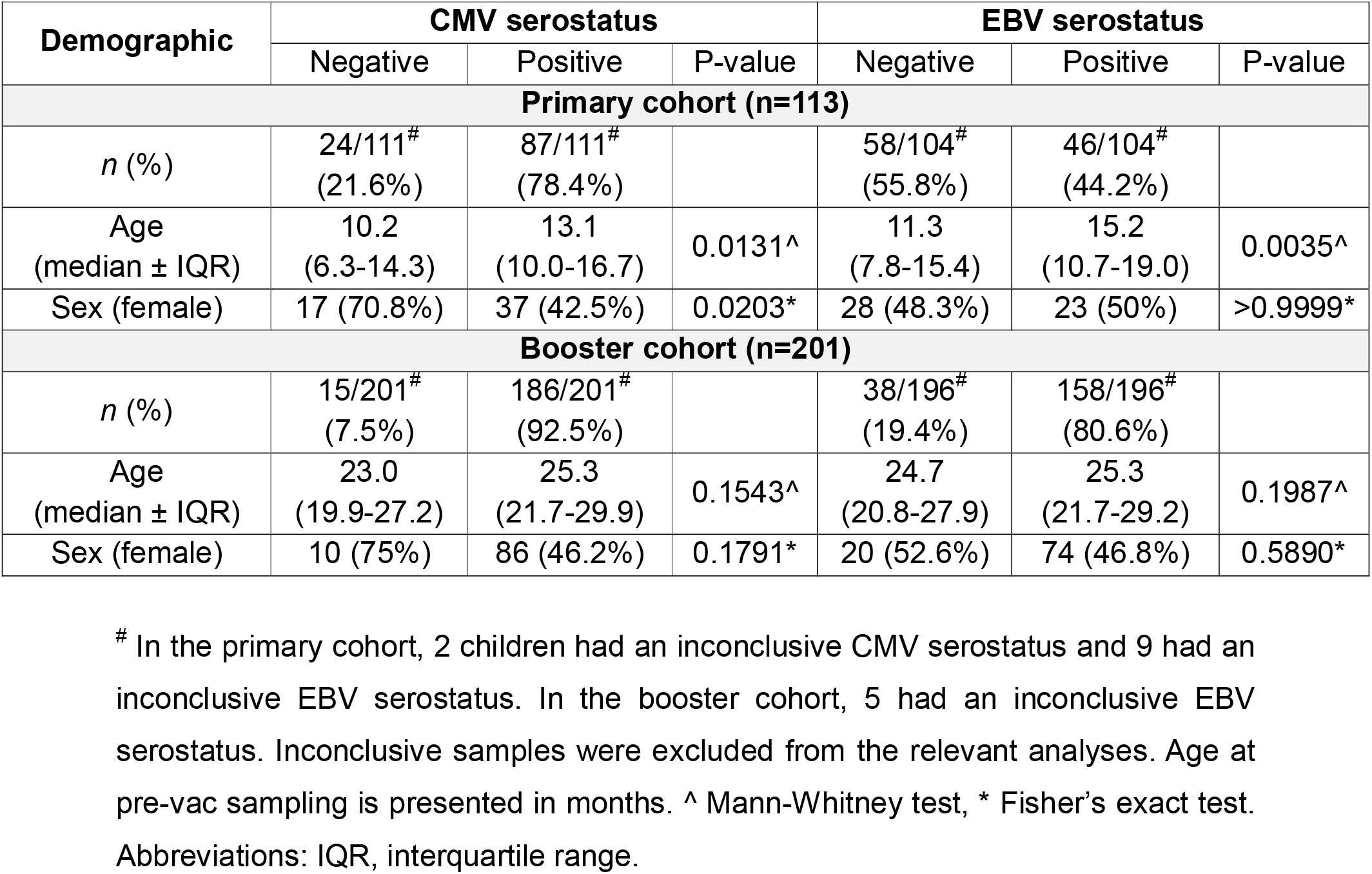
Participant demographics according to CMV or EBV serostatus.

### Impact of CMV and EBV serostatus on RTS,S vaccine-induced antibodies

Post-vac samples were assessed for the induction of antibodies to NANP and C-term antigens following RTS,S vaccination in the primary and booster cohorts. We measured IgG and IgA, the predominant isotypes in serum and have been associated with RTS,S vaccine efficacy^1^, as well as IgM, which has been previously shown to increase following vaccination. All antibody responses had some level of variability among children. In the primary vaccination cohort, the most prominent effect observed was the association of EBV seropositivity (EBV+, n=46; EBV-, n=59) with significantly lower IgG (8% reduction; P=0.0258) and IgA (27% reduction; P=0.0311) antibody isotypes to NANP (Figure 1A). EBV seropositivity was also associated with lower IgG to C-term, although this had weak statistical significance (12% reduction; P=0.0682). In contrast, CMV serostatus had no association with any antibody response expect for IgG to C-term, which was modestly higher in CMV+ children (n=24) than CMV-children (n=88; 12% increase; P=0.0121; Figure 1B).

**Figure 1.**
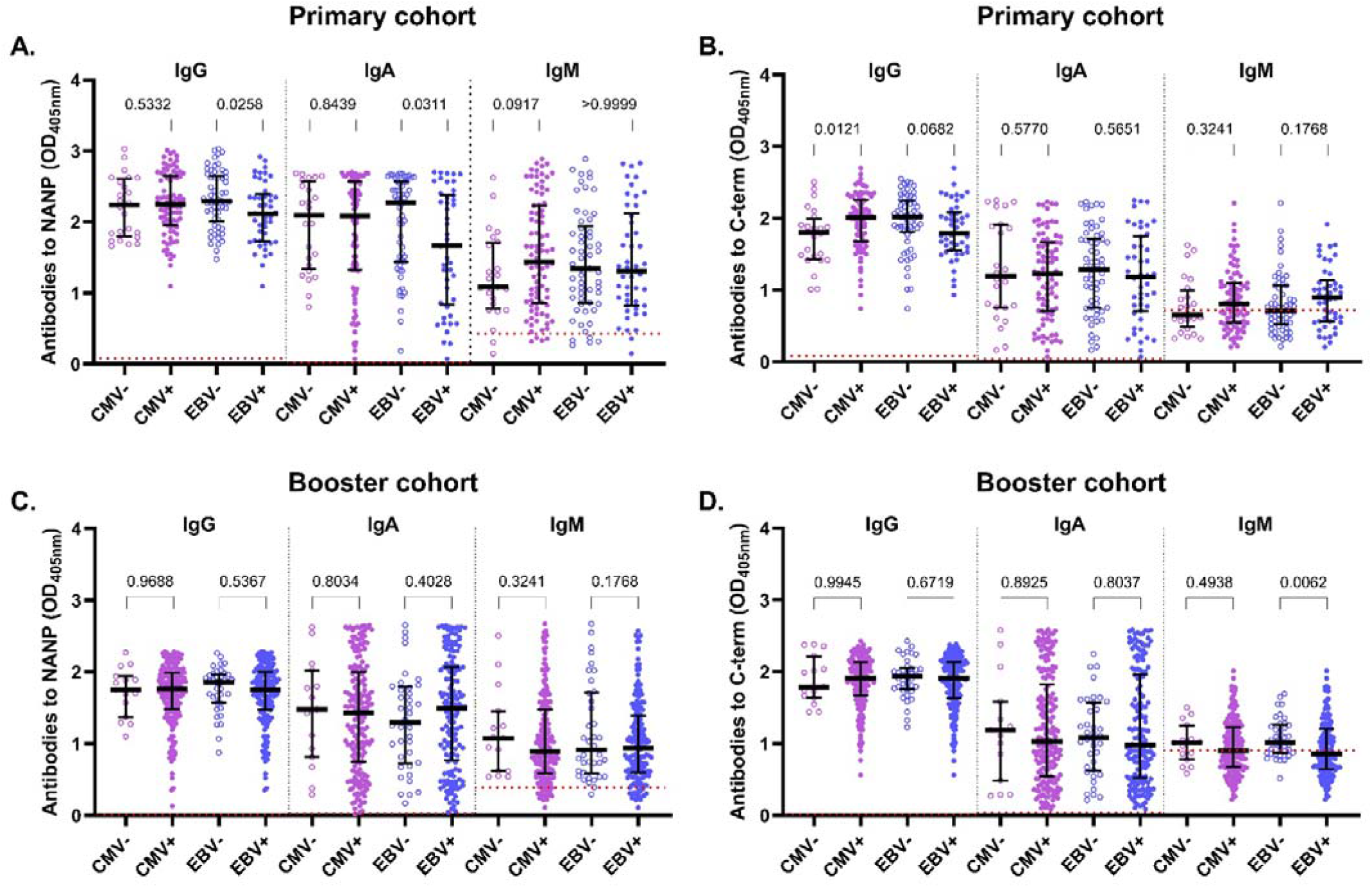
Impact of CMV or EBV serostatus on RTS,S induced IgG, IgA, and IgM antibodies to NANP and the C-term. Children completed the primary (primary cohort) or booster (booster cohort) RTS,S vaccine regimens. Samples collected pre-vac were used to determine baseline viral serostatus, and samples collected one-month post-vac were used to assess vaccine-induced antibodies. Top panels show the induction of IgG, IgA, and IgM to (A) NANP and (B) C-term in the primary cohort among CMV-(open purple circle, n=24), CMV+ (closed purple circle, n=87), EBV-(open blue circle, n=58), and EBV+ (closed blue circle, n=46) children. Bottom panels show the magnitude of IgG, IgA, and IgM to (C) NANP and (D) C-term in the booster cohort among CMV-(open purple circle, n=15), CMV+ (closed purple circle, n=186), EBV-(open blue circle, n=38), and EBV+ (closed blue circle, n=158) children. Lines show median ± interquartile range (IQR). Mann-Whitney Test was used to compare + and - serostatus groups. Red dotted line indicates the seropositivity cut-off calculated as the median of the unvaccinated control group.

In the booster cohort, neither CMV nor EBV serostatus was associated with antibody magnitude following booster vaccination (Figure 1C and 1D). The only significant, though marginal, trend was that IgM isotype to C-term was lower in EBV+ children (n=158) than EBV-children (n=38; 16% reduction; P=0.0062). However, the biological relevance of this finding is limited given that only 50% of the children had detectable IgM above the seropositivity cut-off. We confirmed these associations using multiple linear regression analyses with adjustment for age and sex (Supplementary Table 3). This demonstrated that while EBV seropositivity had negatively impacted IgG and IgA to NANP following primary vaccination, it could explain only ∼5-10% of the variation in antibody level.

## Discussion

This brief report presents the first investigation of the potential impact of CMV and EBV on RTS,S malaria vaccine-induced antibody responses in young children in West Africa. We evaluated two cohorts of children who had samples collected at the time of primary (in 2017) or booster (in 2018) vaccination; therefore, the booster cohort was ∼12 months older than the primary cohort. Seropositivity for CMV or EBV were moderate in the primary cohort and increased in the booster cohort such that >80% of children were seropositive for at least one virus at this later age. This is in line with previously reported data that show most individuals are exposed to these infections in the first year of their life^2,3^. It is important to note that the commercial kits used are unable to distinguish active versus latent viral infection, which could be valuable in future studies.

EBV seropositivity was significantly associated with reduced IgG and IgA to NANP in the primary cohort, which may be important as these antibodies have been correlated with RTS,S vaccine efficacy^1,13^. Notably, this reduction in IgA response (by 26%) was the most substantial reduction observed in our study. This effect was only evident for antibody responses to NANP, but not to C-term. While we did not explore the mechanisms by which EBV reduces RTS,S-induced antibodies, others have shown that EBV transcription factors can suppress B cell differentiation into antibody-secreting cells *ex vivo*, potentially resulting in less serum antibodies and influencing antibody isotype switching^14^.

CMV seropositivity did not negatively impact vaccine-induced antibodies. The only significant association found was that CMV+ children had higher IgG to C-term compared to CMV-children in the primary cohort, but the effect size was marginal and was not consistently observed in the booster cohort. These findings confirm that of other childhood vaccines. For example, Gambian children of a similar age group exhibited no differences in antibody responses to tetanus, *Haemophilus influenzae* type B, measles, or meningococcus vaccinations according to CMV serostatus^6,12^.

Overall, we provide the first evidence that seropositivity for EBV, but not for CMV, is negatively associated with protective antibody responses to primary RTS,S vaccination. While significant, regression analyses revealed that EBV only partly explained the heterogeneity observed in young African children, suggesting that other environmental or host factors may also play a role. Viral serostatus had no effect on the response to booster vaccination, although this could have been limited by the fact that most children were seropositive by the age of the booster dose administration. This may partly explain why vaccine efficacy for R21, and potentially for RTS,S, is reportedly higher in younger children (∼6 months) who are less likely to have been exposed to EBV than older children (∼2 years)^15^. Further investigation of the potential association between viral serostatus and malaria vaccine efficacy is required. Our findings have important implications for the implementation and effectiveness of RTS,S (and the similar R21 vaccine) in regions where EBV is prevalent and impacting vaccine responses. Our data also support ongoing efforts to develop vaccines against EBV to reduce the negative health consequences of this viral infection and to negate the negative effects of EBV on response to childhood vaccines such as RTS,S.

## Acknowledgements

We thank all children, parents and guardians who participated in this study. We thank all investigators and staff who were involved in conducting the clinical trial, sample collection, and data management.

## Financial support

This work was supported by the National Health and Medical Research Council (NHRMC) of Australia (Investigator Grant 2033320 and Synergy Grant 2018654 to JGB), and the Australian Government Research Training Program Scholarship to AWH. AWH, AT, LK, and JGB are members of the NHMRC Australian Centre for Research Excellence in Malaria Elimination. The Burnet Institute is supported by the Victorian State Government Operational Infrastructure Support Grant and the NHMRC Independent Research Institutes Infrastructure Support Scheme. The original phase III trial (MALARIA-055, NCT00866619) was sponsored by GlaxoSmithKline Biologicals SA, and GSK investigators had major roles in the design and analysis of the original study. All the data collection was conducted by local teams, not GSK. GSK did not sponsor this analysis, but GSK did have the opportunity to review and suggest edits to this manuscript. GSK played no role in designing this analysis of the data. The clinical trial was further funded by the UK Joint Global Health Trials (The Department of Health and Social Care, the Department for International Development, the Global Challenges Research Fund, the Medical Research Council and the Welcome Trust) Grant MR/P006876/1. PATH provided additional funding for this work under its grant from the Gates Foundation [INV-007217]. The conclusions and opinions expressed in this work are those of the author(s) alone and shall not be attributed to the foundation. Under the grant conditions of the Foundation, a Creative Commons Attribution 4.0 License has already been assigned to the Author Accepted Manuscript version that might arise from this submission. Please note works submitted as a preprint have not undergone a peer review process.

## Author contributions

AWH, AT, LK, and JGB wrote the manuscript. AWH performed all experiments and analysis. LK, AT, and JGB provided supervision. JBO, HT, IZ, AD, IS, DC, and BG were involved in the clinical trial.

## Conflict of interest statement

The authors declare no competing interests.

## Notes

### Competing Interest Statement

The authors have declared no competing interest.

## References

1. Kurtovic, L. et al. Antibody mechanisms of protection against malaria in RTS,S-vaccinated children: a post-hoc serological analysis of phase 2 trial. The Lancet Microbe 5, (2024).

2. Bates, M. & Brantsaeter, A. B. Human cytomegalovirus (CMV) in Africa: a neglected but important pathogen. J Virus Erad 2, 136–142.

3. Piriou, E. et al. Early age at time of primary Epstein-Barr virus infection results in poorly controlled viral infection in infants from Western Kenya: clues to the etiology of endemic Burkitt lymphoma. J Infect Dis 205, 906–913 (2012).

4. Silva, J. de M., Alves, C. E. de C. & Pontes, G. S. Epstein-Barr virus: the mastermind of immune chaos. Front. Immunol. 15, (2024).

5. Brodin, P. et al. Variation in the human immune system is largely driven by non-heritable influences. Cell 160, 37–47 (2015).

6. Holder, B. et al. Epstein-Barr Virus but Not Cytomegalovirus Is Associated with Reduced Vaccine Antibody Responses in Gambian Infants. PLoS One 5, e14013 (2010).

7. Mukhiya, R. et al. Heterogeneity of the human immune response to malaria infection and vaccination driven by latent cytomegalovirus infection. eBioMedicine 109, (2024).

8. Frasca, D., Diaz, A., Romero, M., Landin, A. M. & Blomberg, B. B. Cytomegalovirus (CMV) seropositivity decreases B cell responses to the influenza vaccine. Vaccine 33, 1433–1439 (2015).

9. Chandramohan, D. et al. Seasonal Malaria Vaccination with or without Seasonal Malaria Chemoprevention. N Engl J Med 385, 1005–1017 (2021).

10. Ali, M. S. et al. The anti-circumsporozoite antibody response to repeated, seasonal booster doses of the malaria vaccine RTS,S/AS01E. npj Vaccines 10, 1–11 (2025).

11. Kurtovic, L. et al. Induction and decay of functional complement-fixing antibodies by the RTS,S malaria vaccine in children, and a negative impact of malaria exposure. BMC Medicine 17, 45 (2019).

12. Miles, D. J. C. et al. Cytomegalovirus infection induces T-cell differentiation without impairing antigen-specific responses in Gambian infants. Immunology 124, 388 (2008).

13. Dobaño, C. et al. Concentration and avidity of antibodies to different circumsporozoite epitopes correlate with RTS,S/AS01E malaria vaccine efficacy. Nat Commun 10, 2174 (2019).

14. Styles, C. T. et al. EBV epigenetically suppresses the B cell-to-plasma cell differentiation pathway while establishing long-term latency. PLoS Biol 15, e2001992 (2017).

15. Datoo, M. S. et al. Safety and efficacy of malaria vaccine candidate R21/Matrix-M in African children: a multicentre, double-blind, randomised, phase 3 trial. The Lancet 403, 533–544 (2024).

