## Supplementary Figure 1; Supplementary Table 1;Supplementary Table 2; Supplementary Table 3 for "Epstein-Barr virus influences the induction of protective antibody responses in young African children vaccinated with the RTS,S malaria vaccine"

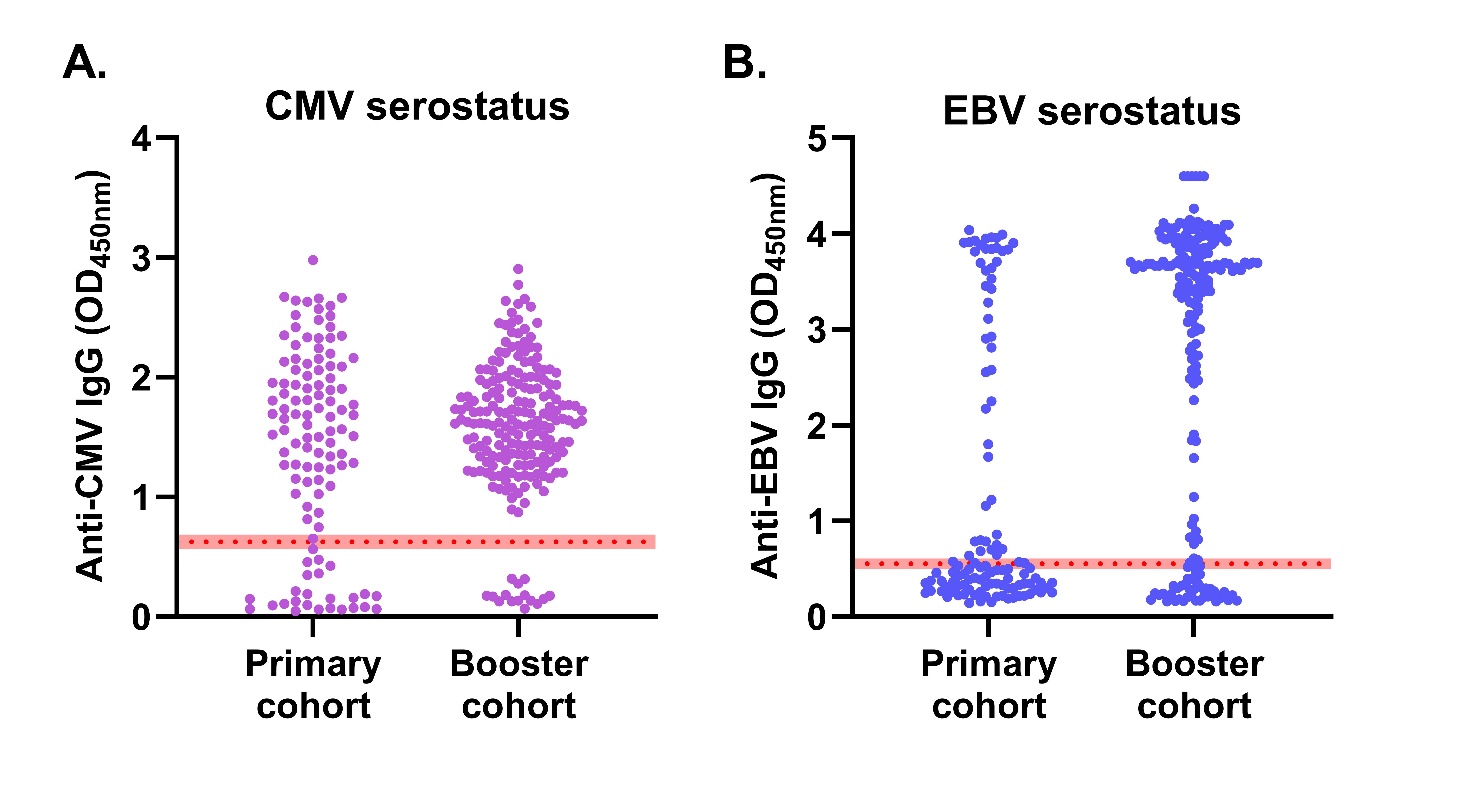

**Supplementary Figure 1. CMV and EBV serostatus of all participants.** Pre-vac samples from children in the primary (n=113) and booster (n=201) cohorts were tested for (**A**) anti-CMV and (**B**) anti-EBV IgG antibodies using kits as per manufacturer’s instructions. The red dotted lines, including the ±10% shown in shaded red, indicate the seropositivity cut-off. In the primary cohort, 2 children had an inconclusive CMV serostatus and 9 had an inconclusive EBV serostatus. In the booster cohort, 5 had an inconclusive EBV serostatus.

**Supplementary Table 1.** **Participant demographics according to CMV/EBV double negative and double positive serostatus.**

| **Demographic** | **CMV-/EBV-** | **CMV+/EBV+** | **P-value** |
| --- | --- | --- | --- |
| **Primary cohort (n=113)** | | | |
| *n* (%) | 18/102^#^ (17.6%) | 40/102^#^ (39.2%) |  |
| Age (median ± IQR) | 10.1 (6.0-13.9) | 15.1 (10.9-17.9) | 0.0005^ |
| Sex (female) | 13 (72.2%) | 19 (47.5%) | 0.0951* |
| **Booster cohort (n=201)** | | | |
| *n* (%) | 6/196^#^ (3.0%) | 149^#^ (76.0%) |  |
| Age (median ± IQR) | 20.5 (19.5-23.8) | 25.2 (21.7-29.3) | 0.0174^ |
| Sex (female) | 4 (66.7%) | 68 (45.6%) | 0.4172* |

^#^ In the primary cohort, 2 children had an inconclusive CMV serostatus and 9 had an inconclusive EBV serostatus. In the booster cohort, 5 had an inconclusive EBV serostatus. Inconclusive samples were excluded from the relevant analyses. Age at pre-vac sampling is presented in months. ^ Mann-Whitney test, * Fisher’s exact test. Abbreviations: IQR, interquartile range.

**Supplementary Table 2. Participant demographics and viral serostatus by study site.**

| **Demographic** | **Burkina Faso** | **Mali** | **P-value** |
| --- | --- | --- | --- |
| **Primary cohort (n=113)** | | | |
| *n* (%) | 77 (68.1%) | 36 (31.9%) |  |
| Age (median ± IQR) | 12.5 (9.1-16.4) | 13.3 (8.1-16.5) | 0.6935^ |
| Sex (female) | 36 (46.8%) | 18 (50.0%) | 0.8405* |
| CMV seropositive | 61 (79.2%) | 26 (72.2%) | 0.8042* |
| EBV seropositive | 33 (42.9%) | 13 (36.1%) | 0.4036* |
| **Booster cohort (n=201)** | | | |
| *n* (%) | 123 | 78 |  |
| Age (median ± IQR) | 24.6 (20.7-28.6) | 25.9 (22.9-28.9) | 0.3836^ |
| Sex (female) | 56 (45.6%) | 40 (51.3%) | 0.4701^ |
| CMV seropositive | 111 (90.2%) | 75 (96.1%) | 0.1694* |
| EBV seropositive | 93 (75.6%) | 65 (83.3%) | 0.0983* |

Age at pre-vac sampling is presented in months. ^ Mann-Whitney test, * Fisher’s exact test. Abbreviations: IQR, interquartile range.

| **Cohort** | **Serostatus** | **Vaccine antigen** | **Antibody** | **Unadjusted** | | | | **Adjusted*** | | | |
| --- | --- | --- | --- | --- | --- | --- | --- | --- | --- | --- | --- |
|  |  |  |  | Coefficient | SE | P | R^2^ | Coefficient | SE | P | R^2^ |
| **Primary**  (n=113) | CMV+ | NANP | IgG | 0.0474 | 0.0993 | 0.6340 | 0.0021 | 0.0930 | 0.1041 | 0.3739 | 0.0392 |
|  |  |  | IgA | -0.1018 | 0.1745 | 0.5609 | 0.0031 | -0.0577 | 0.1859 | 0.7567 | 0.0091 |
|  |  |  | IgM | 0.3197 | 0.1697 | 0.0622 | 0.0315 | 0.3129 | 0.1798 | 0.0848 | 0.0474 |
|  |  | C-term | IgG | 0.2126 | 0.0904 | **0.0205** | 0.0483 | 0.2383 | 0.0949 | **0.0135** | 0.0810 |
|  |  |  | IgA | -0.0841 | 0.1408 | 0.5517 | 0.0033 | -0.0366 | 0.1497 | 0.8072 | 0.0131 |
|  |  |  | IgM | 0.0931 | 0.0956 | 0.3324 | 0.0086 | 0.0848 | 0.0971 | 0.3845 | 0.1051 |
|  | EBV+ | NANP | IgG | -0.2011 | 0.0819 | **0.0158** | 0.0557 | -0.1674 | 0.0854 | 0.0529 | 0.0741 |
|  |  |  | IgA | -0.3573 | 0.1452 | **0.0156** | 0.0560 | -0.3808 | 0.1507 | **0.0131** | 0.0837 |
|  |  |  | IgM | 0.0152 | 0.1454 | 0.9171 | 0.0001 | -0.0526 | 0.1507 | 0.7277 | 0.0306 |
|  |  | C-term | IgG | -0.1160 | 0.0785 | 0.1429 | 0.0209 | -0.0970 | 0.0821 | 0.2404 | 0.0338 |
|  |  |  | IgA | -0.0815 | 0.1190 | 0.4951 | 0.0046 | -0.0605 | 0.1245 | 0.6282 | 0.0169 |
|  |  |  | IgM | 0.1015 | 0.0825 | 0.2215 | 0.0146 | 0.0412 | 0.0828 | 0.6196 | 0.1048 |
| **Booster** (n=201) | CMV+ | NANP | IgG | -0.0310 | 0.1134 | 0.7851 | 0.0004 | -0.0305 | 0.1148 | 0.7907 | 0.0054 |
|  |  |  | IgA | -0.0561 | 0.2021 | 0.7818 | 0.0004 | -0.1209 | 0.2008 | 0.5478 | 0.0437 |
|  |  |  | IgM | -0.1066 | 0.1616 | 0.5102 | 0.0022 | -0.1106 | 0.1590 | 0.4875 | 0.0628 |
|  |  | C-term | IgG | -0.0362 | 0.0981 | 0.7126 | 0.0007 | -0.0315 | 0.0991 | 0.7505 | 0.0104 |
|  |  |  | IgA | 0.0031 | 0.1987 | 0.9874 | 0.0000 | -0.0113 | 0.1989 | 0.9548 | 0.0288 |
|  |  |  | IgM | -0.0466 | 0.0985 | 0.6371 | 0.0011 | -0.0307 | 0.0960 | 0.7497 | 0.0800 |
|  | EBV+ | NANP | IgG | -0.0737 | 0.0737 | 0.3191 | 0.0051 | -0.0686 | 0.0743 | 0.3570 | 0.0100 |
|  |  |  | IgA | 0.1218 | 0.1361 | 0.3718 | 0.0041 | 0.0862 | 0.1350 | 0.5240 | 0.0403 |
|  |  |  | IgM | -0.1318 | 0.1070 | 0.2196 | 0.0078 | -0.1478 | 0.1049 | 0.1604 | 0.0670 |
|  |  | C-term | IgG | -0.0693 | 0.0657 | 0.2929 | 0.0057 | -0.0636 | 0.0661 | 0.3376 | 0.0144 |
|  |  |  | IgA | 0.1117 | 0.1331 | 0.4025 | 0.0036 | 0.0959 | 0.1323 | 0.4695 | 0.0361 |
|  |  |  | IgM | -0.1458 | 0.0656 | **0.0274** | 0.0248 | -0.1495 | 0.0634 | **0.0194** | 0.1088 |

**Supplementary Table 3. Multiple linear regression exploring the impact of CMV or EBV status on antibody responses in the primary and booster cohorts, adjusting for ag and sex.**

Seronegative children served as the reference group. *Adjusted for age and sex.
